# Non-invasive finger sweat analysis provides distribution-related information for dexamphetamine and zolpidem in humans

**DOI:** 10.64898/2026.11.19.26368478

**Authors:** Michael Wolf, Veronika Pfoser-Poschacher, Daniel Wasinger, Gregor Hahn, Samuel M. Meier-Menches, Michael Wolzt, Christopher Gerner

## Abstract

Eccrine sweat reportedly contains information about drug distribution, but its collection is often lengthy and sophisticated. Finger sweat is collected within 3 minutes by standardized extraction of fingertips, simple, and non-invasive. We previously demonstrated that finger sweat indicates drug intake, motivating us to assess its distribution-related information. Thus, two well characterized drugs with contrasting distribution characteristics were chosen for the parallel analysis of finger sweat and plasma samples. Forty participants received a single-dose of 30 mg dexamphetamine hemisulfate or 5 mg zolpidem tartrate (n = 20 per group). Matched finger sweat and plasma samples collected at baseline and 3 and 8 h post-dose were analyzed by mass spectrometry hyphenated with liquid chromatography. Assuming 1 microliter sweat per sample, we calculated apparent finger sweat concentrations and finger sweat-to-total-plasma concentration ratios. All 40 corresponding post-dose samples for each drug were positive and all 80 expected-negative samples were negative. Finger sweat-to-total-plasma concentration ratio was approximately 4-fold higher for dexamphetamine compared with zolpidem at 3 h (*p* = 9.9 10E-6) and 8 h (*p* = 2.3 10E-5) and approximately doubled from 3 to 8 h for both dexamphetamine (*p* = 0.013) and zolpidem (*p* = 6.7 10E-5). The higher finger sweat-to-total-plasma concentration ratio values for dexamphetamine are consistent with its lower plasma protein binding and larger apparent volume of distribution, providing initial evidence that finger sweat may contain distribution-related information. The time-dependent finger sweat-to-total-plasma concentration ratio increases are compatible with slow drug equilibration and retention within the stratum corneum. The present findings warrant further investigation of finger sweat as an easily accessible sample matrix to assess distribution-related drug characteristics.

## 1 Introduction

Translational pharmacology aims to predict drug response by linking unbound plasma exposure to drug concentrations at or near sites of action (1, 2). In physiologically based pharmacokinetic/pharmacodynamic models, tissue exposure is commonly estimated using tissue-to-plasma partitioning coefficients determined in animal models together with physiological and drug-specific parameters (2-4). These estimates often rely on simplifying assumptions, such as passive distribution, rapid tissue-plasma equilibration, and homogeneous tissue compartments, which may limit prediction accuracy in vivo (1, 4). Direct assessment of tissue drug exposure or partitioning in humans remains challenging and may vary between and within individuals because of differences in membrane permeability, perfusion, tissue binding, pH-dependent partitioning, transporter activity, or pathophysiological conditions (4-7). Direct measurement of drug concentrations closer to peripheral target sites, such as tissue interstitial fluid (ISF), could improve the interpretation of plasma pharmacokinetics and exposure-response relationships (8, 9).

Techniques such as microdialysis (8, 9), tissue biopsy (10), cerebrospinal fluid sampling (11), and molecular imaging (12) can provide valuable information in this context. However, these methods are often invasive, technically demanding, unsuitable for repeated sampling, or prone to substantial variability. In contrast, sweat-based sampling may offer non-invasive, simple, and repeatable alternatives to acquire information about drug distribution and tissue penetration (13). Eccrine sweat is derived from dermal ISF surrounding sweat glands (14) and may serve as a proxy for drug penetration into ISF, as has been recently demonstrated with antibiotics (15). Therefore, drug quantification in eccrine sweat may be valuable to study drug distribution.

The present study used an understudied sampling approach for sweat analysis, in which pre-wetted paper is pressed between the thumb and index finger for 1 min. The resulting extracted sample material, termed finger sweat (FiS), differs from conventional sweat samples by containing freshly secreted insensible eccrine sweat as well as residual components retained in the stratum corneum (16). A FiS sample therefore represents a standardized aqueous extract of the skin and sweat compartments that contains eccrine sweat, but with unique characteristics. Compared with the collection of pure eccrine sweat, this pragmatic approach improves time-dependent analyte recovery due to its surprisingly extensive metabolomic coverage (17, 18). Importantly, this simple sample collection can be performed without trained personnel (17-19). Recent studies demonstrated the feasibility of this approach to determine various drugs in the fields of oncology, psychiatry, and forensic toxicology (20-22). However, it remains unclear whether FiS contains quantitative information related to drug distribution and tissue penetration.

This proof-of-concept study investigated matched FiS and blood samples with regard to two well-characterized drugs with contrasting distribution behaviors at baseline and 3 and 8 h post-dose (n = 20 per drug). This study design allows us to address the following questions: 1) Are both drugs detectable in FiS samples? 2) Does the ratio between FiS and total plasma concentrations, defined as R_FiS/P_ = [drug]_FiS_ / [drug]_plasma_, agree with the expected distribution characteristics of both drugs? 3) Is it possible to obtain relevant information from FiS analysis by comparing the two time points? We hypothesized that dexamphetamine would show a higher R_FiS/P_ than zolpidem, consistent with its lower protein binding and greater apparent peripheral distribution (2, 23).

## 2 Materials and Methods

### 2.1 Study design

In the frame of a registered clinical study (24), two randomized groups of 20 healthy males were orally administered either 30 mg dexamphetamine hemisulfate (equivalent to 22 mg dexamphetamine; Attentin®; MEDICE) or 5 mg zolpidem tartrate (equivalent to 4 mg zolpidem; Zolpidem HEXAL®; Hexal AG) in a parallel-group design. FiS and EDTA-anticoagulated venous blood samples were collected at baseline and 3 h and 8 h post-dose, resulting in a total of 120 samples per matrix. Blood samples were centrifuged at 2’000 RCF at 4 °C for 10 min and plasma was aliquoted into polypropylene cryovials. Plasma and FiS samples were transported and stored at 80 °C.

The study protocol obtained approval from the Ethics Committee of the Medical University of Vienna (EK Nr: 1106/2022) and from the Federal Office for Safety in Health Care (BASG; Reference Nr: 10070499). The trial is registered under the EudraCT identifier number 2021-005381-17. Informed consent was obtained from all participants.

### 2.2 Finger sweat sample collection

Paper discs consisting of circularly punched precision wipes (KIMTECH Science precision wipes 7552; Kimberly-Clark Professional) with a diameter of ½ inch were pre-wetted with 3 µL H_2_O and stored in 0.5 mL safe-lock tubes (Eppendorf) until use. The skin/sweat compartment from the fingertips was sampled by the following procedure: hands were briefly rinsed with tap water and dried using paper towels. After a 1 min delay, a paper disc was placed between index finger and thumb using tweezers and firm pressure was applied for 1 min. Finally, the paper disc was placed back into the original 0.5 mL tube, resulting in a total sampling time of less than 3 minutes.

### 2.3 Finger sweat sample extraction

Sample extraction was performed in a randomized order. Each sample-containing tube received 70 µL of a mixture containing water (LC-MS grade; VWR Chemicals), methanol (MeOH; LC-MS grade; Honeywell), and formic acid (FA; LC-MS grade; VWR Chemicals) at a volume ratio of 97/3/0.2, respectively. The paper disc was pressed to the bottom of the tube using the pipette tip. After 30 s in an ultrasonic bath (100%, 25 °C; SONOREX DIGITAL 10P; BANDELIN electronic), the supernatant was transferred into HPLC-vials equipped with 200 µL glass inserts (Macherey-Nagel). Process blanks contained a clean paper disc and were subjected to the same procedure with the purpose of quality control. Of each sample, 10 µL were injected for Ultra-high-performance liquid chromatography–high-resolution mass spectrometry (UHPLC-HRMS) analysis directly following extraction.

### 2.4 Plasma extraction

Plasma samples were thawed on ice and 40 µL was transferred into 1.5 mL safe-lock tubes (Eppendorf). Samples were diluted 1:5 with MeOH and incubated over night at -20 °C to precipitate proteins, followed by a solid phase extraction (SPE) using Strata-X polymeric reversed-phase cartridges (33 µm particle size; 10 mg; Phenomenex).

After centrifugation for 10 min at 4 °C and 12’000 RCF, 25 µL of each supernatant was loaded onto MeOH-conditioned SPE cartridges. Analytes were eluted into 1.5 mL HPLC-vials using 300 µL MeOH containing 20 mM ammonium carbonate (Fluka), and the eluates were evaporated under nitrogen. The residues were reconstituted in 200 µL H_2_O/MeOH/FA (97/3/0.2; v/v/v) and transferred into 200 µL glass inserts. For UHPLC-HRMS analysis, 5 µL was injected per sample directly following extraction, representing 0.125 µL EDTA-plasma on column.

### 2.5 Sample analysis

FiS and plasma samples were measured at identical conditions using an Infinity 1290 UHPLC coupled to an Orbitrap Exploris 480 mass spectrometer. A Kinetex XB-C18 column (1.7 µm, 100 × 2.1 mm; Phenomenex Inc.) was used for chromatographic separation at 40 °C. Mobile phase A consisted of H_2_O and mobile phase B of MeOH, both containing 0.2% FA. Gradient elution was performed at a flowrate of 0.5 mL min^-1^ and started with an isocratic hold at 3% B for 0.2 min, followed by a linear ramp to 8% B until 1.3 min, and reached 100% B by 6.2 min. A 0.3 min wash out phase at 100% B was followed by an injection of 10 µL methanol with a 0.6 min long wash at 95% B. Finally, the column was equilibrated at starting conditions for 2.2 minutes.

Electrospray-ionization operated at 3200 V in positive-mode with a sheath gas, aux gas, and sweep gas of 60, 9, and 1 AU, respectively. The ion transfer tube was set to 325 °C, auxiliary gas to 340 °C, RF lens to 50%, AGC-target to 100%, resolution to 90’000, EASY-IC^TM^ to scan-to-scan, and maximum injection time to 182 ms. The instrument measured protonated dexamphetamine (m/z = 136.1121) and zolpidem (m/z = 308.1757) in tSIM mode at a retention time of 2.18 and 3.38 min, respectively. The system was controlled using Xcalibur (ver. 4.6.67.17), acquired data was evaluated and integrated using Skyline (ver. 25.1.0.237), and statistical analysis was performed using R (ver. 4.5.2) (25, 26).

### 2.6 Quantification and partial validation

Tablet-derived standards for quantification and partial validation were prepared by homogenizing Attentin (10 mg dexamphetamine hemisulfate; MEDICE) and Zolpidem HEXAL (10 mg zolpidem tartrate; Hexal AG) tablets and dissolving weighted portions with MeOH.

Blank paper discs were spiked with standards and dried for validating FiS analysis. Plasma analysis validation was performed by spiking standard solutions 1:10 into EDTA-anticoagulated reference plasma aliquots. To calculate apparent concentrations in FiS samples, we assumed that each sample collected contains 1 µL sweat, as the index finger has a reported sweat rate between 93–275 nL min^-1^ cm^-2^ in a comfortable room (27). Assuming equal sweat rates on thumbs, the sweat volume per sample approximates (0.635 cm)^2^ × π × 2 fingers × 2 min × sweat rate = 471–1393 nL sweat per sample. We additionally assumed full evaporation of pre-wetting solution and sweat residue pre-extraction.

Calibration curves were prepared in triplicates by 1:2 serial dilutions at 9 calibration points, covering 7–1792 ng mL^-1^ (dexamphetamine) and 0.7–179.2 ng mL^-1^ (zolpidem) in FiS and 1–256 ng mL^-1^ (dexamphetamine) and 0.4–102.4 ng mL^-1^ (zolpidem) in plasma. Calibration points with back calculated concentrations deviating > ±15% from their nominal concentrations were removed. Precision, accuracy, and recovery were assessed using mid-level QCs at respective concentrations to calibrant point 5 (28). Precision and accuracy were determined using 6 independent replicates covering variance from sample extraction and UHPLC-HRMS analysis. Recovery was calculated by the mean signal of 6 replicates spiked pre-extraction divided by the mean signal of 6 replicates spiked post extraction.

## 3 Results

### 3.1 Cohort characteristics and validation summary

A total of 40 healthy male Caucasian subjects were randomized to orally ingest medication containing either 22 mg of dexamphetamine or 4 mg of zolpidem. Participants were between 20 and 43 years (27 ± 5 years) with a body mass index (BMI) between 18.7 and 27.2 kg m^-2^ (22.8 ± 2.4 kg m^-2^) and a body weight between 58.0 and 89.3 kg (74.0 ± 9.0 kg). All FiS and EDTA-anticoagulated plasma samples at baseline and 3 and 8 h post-dose were successfully collected, measured *via* tSIM using an Orbitrap Exploris 480, and processed, resulting in a total of 240 samples.

External calibration curves for dexamphetamine and zolpidem were prepared by spiking either blank paper discs for FiS or reference plasma at 9 concentrations before extraction. Each of the four calibration curves was weighted 1/x and passed accuracy and precision criteria for at least 6 calibration points according to FDA M10 guidelines (28). Each of the four mid-level QCs represents the respective concentration at calibration point 5 and passed precision, recovery, and carry-over requirements (28). Quantitative data of the partial validation is summarized in table 1.

**Table 1.**
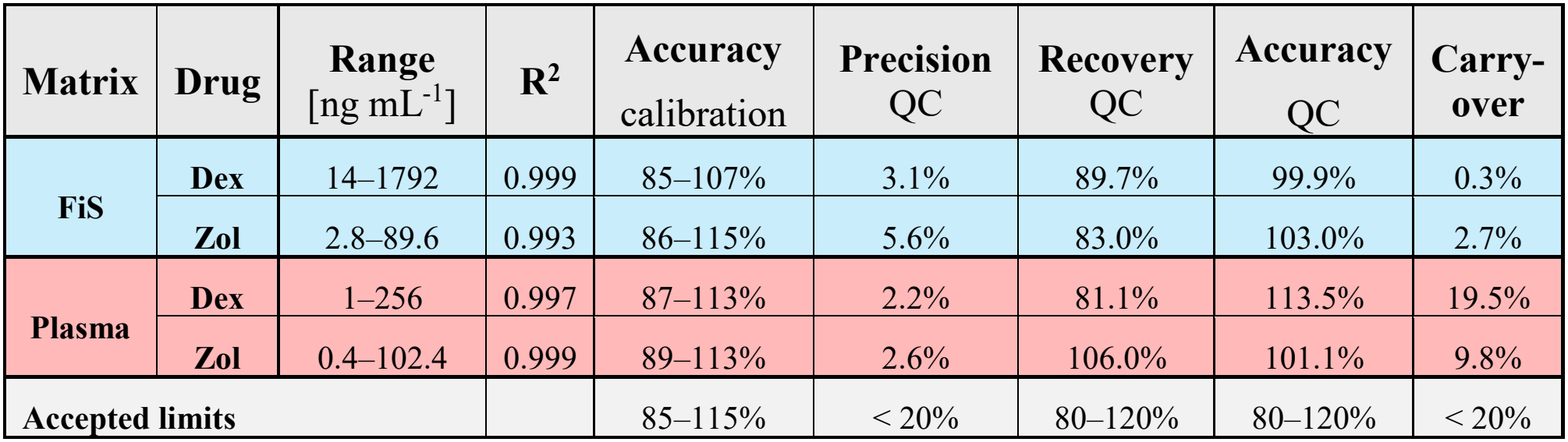
Calibration range and summary of the partial validation of dexamphetamine and zolpidem in FiS and plasma. The lowest concentration in **Range** represents the LOQ. **Accuracy** is reported for the range for all calibration measurements and the mid-level QC. **Precision** and **Recovery** were determined using 6 replicates at mid-level QCs. **Carry-over** was determined as the quotient of the blank injection signal after the highest calibration point and LOQ signal. Dex = dexamphetamine. Zol = zolpidem.

In all post-dose samples, the respective drug was successfully detected, and no dexamphetamine or zolpidem was found in any baseline sample or in the cross-group. All quantified samples were within the validated calibration range, except for one FiS sample containing zolpidem, which exceeded the upper limit of quantification (ULOQ).

### 3.2 Absolute drug quantification in finger sweat

Apparent concentrations in FiS were determined by assuming a sweat volume of 1 µL per sample as described in the method section and are illustrated alongside plasma concentrations in figure 1. Absolute drug amounts in all positive FiS samples ranged from 42–1478 pg per sample for dexamphetamine and 3–156 pg per sample for zolpidem, corresponding to apparent concentrations of approximately 1.9–67 ng mL^-1^ mg _(drug)_ ^-1^ and 0.7–39 ng mL^-1^ mg_(drug)_ ^-1^, respectively.

**Figure 1.**
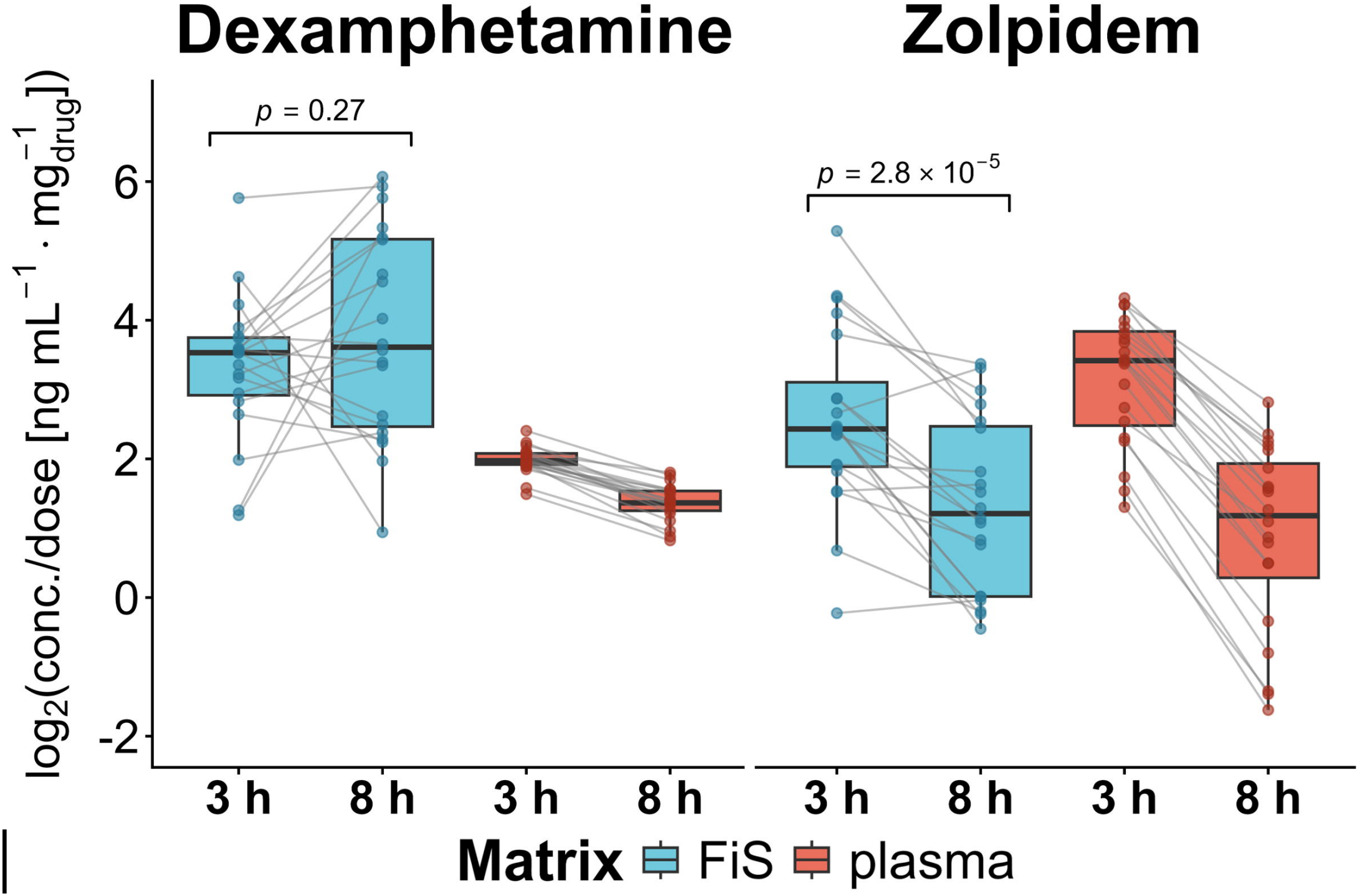
Log_2_-transformed ng drug per mL FiS or plasma normalized to drug dose 3 and 8 h post-dose for dexamphetamine (left) and zolpidem (right). A sweat volume of 1 µL sweat per FiS sample was assumed. Time-dependent R_FiS/P_ differences in drug concentrations in FiS between 3 and 8 h post-dose for each drug were tested with a double-sided paired t-test. Normal distribution was assessed by Shapiro-Wilk test and visual inspection of histograms and homoscedasticity was assessed using an F-test.

**Figure 2.**
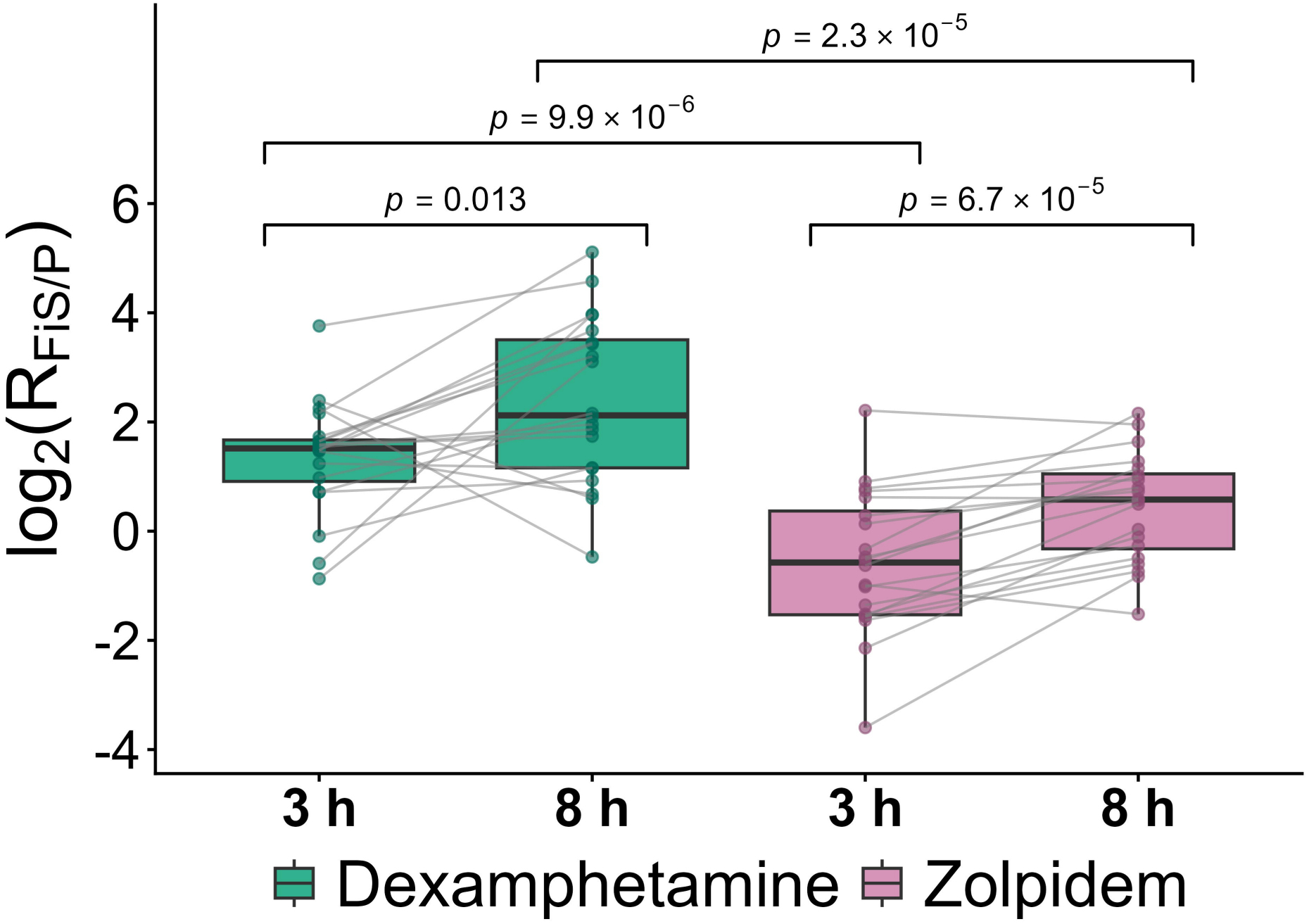
Log_2_-transformed R_FiS/P_ boxplots for dexamphetamine and zolpidem 3 and 8 h post-dose with lines connecting individuals. Differences between dexamphetamine and zolpidem were tested with a double-sided Welch t-test. Time-dependent differences between 3 and 8 h post-dose for each drug was tested with a double-sided paired t-test. Normal distribution was assessed by Shapiro-Wilk test and visual inspection of histograms and equality of variances was assessed using an F-test.

The dexamphetamine group exhibited no differences in FiS drug concentrations between 3 and 8 hours (double-sided paired t-test *p* = 0.27), but significantly higher variances in FiS than plasma at 3 h (*F* = 26.9, *p* = 1.3 × 10^-9^) and 8 h (*F* = 31.0, *p* = 3.7 × 10^-10^). In contrast, the zolpidem group exhibited a significant decrease in FiS concentrations from 3 to 8 hours (double-sided paired t-test *p* = 2.8 × 10^-5^), but had no significant differences in variances at 3 h (*F* = 2.0, *p* = 0.14) and 8 h (*F* = 0.9, *p* = 0.76).

### 3.3 Concentration ratios between FiS and plasma

According to the hypothesis, the drug distribution between plasma and dermal ISF is at least in part reflected by the ratio of apparent FiS and total plasma concentrations R_FiS/P_, which is predicted to increase for drugs with low protein binding and a high V_d_ (2). The arithmetic mean of R_FiS/P_ was 3.2 at 3 h and 8.6 at 8 h in the dexamphetamine group and 1.0 at 3 h and 1.7 at 8 h in the zolpidem group. Consistent with the hypothesis, R_FiS/P_ was significantly higher for dexamphetamine than for zolpidem after 3 h (geometric mean ratio, 3.8; 95% CI, 2.24–6.43; double-sided Welch’s *p* = 9.9 × 10^-6^) and 8 h (geometric mean ratio, 3.9; 95% CI, 2.23–6.93; double-sided Welch’s *p* = 2.3 × 10^-5^). In accordance with the suspected time lag from slow tissue equilibration (29), R_FiS/P_ increased significantly within-subject from 3 to 8 h for both dexamphetamine (geometric mean ratio, 2.1; 95% CI, 1.19–3.72; double-sided paired t-test *p* = 0.013) and zolpidem (geometric mean ratio, 2.0; 95% CI, 1.52–2.72; double-sided paired t-test *p* = 6.7 × 10^-5^).

## 4 Discussion

This proof-of-concept study demonstrates that FiS can provide both qualitative and distribution-related information about systemically administered drugs. Dexamphetamine and zolpidem were reliably detected in every expected post-dose FiS sample at 3 and 8 h (n = 40 per drug), whereas all baseline and cross-group samples were negative (n = 80 per drug). Moreover, R_FiS/P_ was consistently higher for dexamphetamine than for zolpidem, agreeing with the prespecified hypothesis based on their contrasting plasma protein binding and V_d_. Finally, R_FiS/P_ significantly increased between 3 and 8 h for both drugs, indicating that repeated FiS sampling can capture time-dependent differences in the relationship between FiS and plasma concentrations. Collectively, these results support FiS as a promising non-invasive sampling matrix to acquire information about adherence and drug distribution.

### 3.4 Evaluation of FiS for adherence monitoring

The primary requirement for adherence monitoring is the reliable discrimination between drug intake and non-intake. Adherence monitoring is particularly attractive for FiS analysis, because sample collection is simple, rapid, non-invasive, and can be performed repeatedly without trained personnel. These characteristics may be useful when conventional blood sampling is impractical or burdensome, such as outpatient follow-up, psychiatric treatment, or pediatric studies.

Within this controlled dataset, both drugs were detected with 100% true-positive and true-negative rate when concentrations above the limit of quantification were considered positive. Drug amounts in FiS ranged from approximately 42–1478 ng mL^-1^ and 3–156 ng mL^-1^ under the 1 µL assumption. While these findings indicate perfect discrimination, the risk of external contamination remains an important consideration for skin-surface-based matrices (14, 30). Additionally, both drugs showed a higher R_FiS/P_ 8 h post-dose compared to 3 h post-dose, indicating a longer detection window in FiS compared with plasma. Previous forensic work assessed a similar sampling approach in deceased individuals and compared it with a standard toxicological assay using tissue samples (22). They suggested that detecting drug metabolite patterns in addition to the parent compound can improve specificity and reduce false-positive interpretation caused by contamination (22). In agreement with our data, they reported several cases where they classified cocaine, heroin, and/or ketamine consumption despite negative toxicology results, indicating a longer detection window (22).

Overall, the present study demonstrates the potential of FiS sampling and analysis for adherence monitoring, although broader clinical use should consider drug-specific validation of detection windows, metabolite patterns, and contamination controls. Interestingly, the high drug concentrations suggest that future targeted applications may not always require ultra-sensitive LC-MS workflows.

### 4.2 FiS-to-plasma ratio as an index of drug distribution

Dexamphetamine has low plasma protein binding of approximately 16% and a relatively high V_d_ of approximately 3–4 L kg^-1^ (31, 32), whereas zolpidem is approximately 92% protein-bound and has a substantially lower V_d_ of approximately 0.54–0.68 L kg^-1^ (33). As only unbound drug in plasma is considered available for distribution into tissues (1, 2), total plasma concentrations were determined to produce a stark contrast in R_FiS/P_ between both drugs. The approximately 4-fold higher geometric mean of R_FiS/P_ for dexamphetamine at both time points is therefore consistent with its greater availability for peripheral distribution. This finding provides initial evidence that FiS may contain drug distribution-related information.

Absolute values of R_FiS/P_, specifically values above one, should be interpreted cautiously, as secretion mechanisms for most small molecules *via* sweat remain largely unknown (14, 34). FiS can be considered a standardized aqueous extract containing insensible eccrine sweat together with drug retained in the stratum corneum (14, 16). Due to the pH decrease from dermal ISF to sweat, ion trapping may affect tissue equilibration and concentrate basic drugs such as dexamphetamine and zolpidem (5, 6, 34-36). In addition, apparent FiS concentrations were calculated assuming a plausible sample volume of 1 µL sweat according to the sampled area (2.53 cm^2^), sampling time (2 min), and reported sweat-rate ranges on the index finger in typical clinical research conditions (93–275 nL min^-1^ cm^-2^)(27). However, this approach does not account for variability introduced by differences in sweat rates. Thus, R_FiS/P_ does not describe a conventional tissue-to-plasma partition coefficient, but an empirical distribution-related index.

The approximately twofold increase in R_FiS/P_ from 3 to 8 h for both drugs may indicate slower disappearance from the sampled skin/sweat compartment compared with plasma. This observation may be explained by the surprisingly slow tissue equilibration between plasma and dermal ISF, which may lead to a time-delay of the pharmacokinetic profile in FiS compared with plasma (29). However, the fenestrated capillaries and relatively high perfusion and innervation at the fingertips should lead to a different ISF composition with a faster turnover compared with other skin locations (29, 37). Furthermore, drug retention in the stratum corneum may also lead to this pattern, because higher drug amounts may be accumulated in the stratum corneum at 8 h compared with 3 h post-dose (16, 30). The distinction of these mechanisms may be enabled by denser post-dose sampling in regular intervals.

### 4.3 Interindividual variability of FiS concentrations

For zolpidem, FiS and plasma variances were not significantly different at 3 h (*F* = 2.0, *p* = 0.14) or 8 h (*F* = 0.9, *p* = 0.76), and FiS values decreased by a factor of 2.4 from 3 to 8 h post-dose (*p* = 2.8 × 10^-5^). Notably, zolpidem has a short half-life and high plasma protein binding compared with dexamphetamine (32, 33, 38). These characteristics may lead to substantial variability in total plasma concentrations at single time points, especially due to interindividual changes in protein binding (2, 23). The variability of unbound plasma exposure, however, is recognized as unaffected by changes in protein binding for highly protein-bound, low-extraction drugs, leading to a lower time-dependent variability (2, 23). Importantly, only the unbound drug enters the ISF and reaches FiS (4, 14), and equilibration between skin ISF and plasma is slow (29). We therefore suspect that the variability in FiS stemming from the sampling process is compensated by the more stable unbound drug concentration to match the time- and protein-binding-dependent variability of zolpidem in total plasma concentrations.

For dexamphetamine, FiS variance was significantly higher than plasma variance at 3 h (*F* = 26.9, *p* = 1.3 × 10^-9^) and 8 h (*F* = 31.0, *p* = 3.7 × 10^-10^), and FiS values increased by a factor of 1.4 from 3 to 8 h post-dose (*p* = 0.27). In comparison with zolpidem, the variability at single time points of dexamphetamine in total plasma concentrations is reduced by its long half-life and low protein binding (32, 38), leading to a much lower deviation as seen in figure 1. In FiS, however, additional variability may be introduced by pH-dependent sweat partitioning of dexamphetamine, which is comparable to the significant influence of urine pH in its renal elimination (35, 39). Furthermore, the variance of dexamphetamine in FiS may be confounded by its stimulating effects, which was observed to decrease skin perfusion and increase sweat rate, particularly on palmar skin (14, 40, 41).

Unlike zolpidem, dexamphetamine concentrations in FiS did not decrease from 3 to 8 hours post-dose. This finding supports the proposed slow equilibration mechanism between plasma and dermal ISF, as dexamphetamine has a plasma Tmax of about 3.3 h (95% CI, 2.7–3.9) (42), which would be delayed in a slowly equilibrating compartment. This already slow equilibration would be additionally slowed down by the decreased perfusion caused by the stimulating drug effects of dexamphetamine (29).

### 4.4 Limitations

First, two drugs were compared at two post-dose time points, limiting mechanistic interpretation and generalizability to other compounds. As sampling was limited to two time points, pharmacokinetics, equilibration time lags, and detection windows could not be determined. Second, FiS is collected by standardized aqueous extraction of the fingertips and may therefore differ from conventional sweat samples. This pragmatic approach seems to increase time-dependent analyte recovery by extracting sweat deposits in the stratum corneum (16), but also introduces susceptibility to external contamination and complicates mechanistic interpretation. The assumed 1 µL sweat volume is plausible for the sampling process, but differences in recovered volume between samples were not accounted for in this study design. However, the reported sweat rate of 93–275 nL min^-1^ cm^-2^ in resting individuals means that the maximum error in collected sweat volume should not exceed 3-fold changes. Third, only FiS and total plasma concentrations were measured, preventing the assessment of the relationship between FiS, dermal ISF, and unbound plasma concentrations. The observed differences between dexamphetamine and zolpidem are consistent with their known distribution and protein binding properties, but the underlying mechanisms cannot be separated: plasma protein binding (2, 23), slow equilibration of dermal ISF (29), pH-dependent ion trapping (5, 35), and retention within the stratum corneum (16) may all contribute to varying degrees.

### 4.5 Conclusions

Overall, the present study shows that the administered drugs dexamphetamine and zolpidem can be detected in FiS with high specificity in this controlled setting. They were collected in sufficient amounts for quantification and distinguished based on their contrasting distribution properties. The observations in the dataset can be readily explained by known physiology, but detailed mechanisms involved in the sampling process remain to be elucidated. Nevertheless, this proof-of-concept study provides the groundwork for improved study designs to further evaluate the potential of this non-invasively collected matrix for adherence monitoring and assessment of drug distribution that may pave the way for pharmacokinetic modelling in humans.

## Data Availability

The raw data is publicly available on the MassIVE repository under the accession number MSV000103849.

https://massive.ucsd.edu

